# Grip strength is associated with retinal and choroidal thickness in type 2 diabetes mellitus patients without retinopathy

**DOI:** 10.1101/2020.01.31.20019885

**Authors:** Zihan Qiu, Wei Wang, Yan Tan, Miao He, Lanhua Wang, Yuting Li, Xia Gong, Wenyong Huang

**Author notes:** Co-correspondence authors. **Corresponding author:** Wenyong Huang, MD&PhD, Professor of Ophthalmology, Vice director of Department of Preventive Ophthalmology, Zhongshan Ophthalmic Center, State Key Laboratory of Ophthalmology, Sun Yat-sen University, 54S.Xianlie Road, Guangzhou, China 510060, Dr. Wei Wang, MD&PhD, Zhongshan Ophthalmic Center, State Key Laboratory of Ophthalmology, Sun Yat-sen University, Guangzhou, China 510060. **Funding:** This research was supported in part by the National Natural Science Foundation of China (81570843; 81530028; 81721003).

## Abstract

**Objective:** To determine the relationship between grip strength and retinal or choroidal thickness in Chinese patients with type 2 diabetes mellitus.

**Design:** Observational study-cross-sectional design.

**Setting and Participants:** The study was conducted among diabetes patients without retinopathy registered in the community health system in Guangzhou, China.

**Measures:** Grip strength was measured twice for each hand with a dynamometer in kilograms (kg). The retinal and choroidal thickness in macular Early Treatment Diabetic Retinopathy Study (ETDRS) sectors were measured by commercial swept-source optical coherence tomography (SS-OCT; DRI OCT-2 Triton; Topcon, Tokyo, Japan).

**Results:** A total of 1,029 patients were included in the study. Both retinal thickness and choroidal thickness increased with the higher quartile of grip strength. Regression analyses indicated that the average retinal and choroidal thickness increased by 0.14 μm (95%CI: 0.03-0.25 μm, P=0.011) and 0.57 μm (95%CI: 0.03-1.11 μm, P=0.037) for each additional kg of grip strength following adjustment for age and gender. Further adjustments were made for axial length, HbA1c, length of time the patient had diabetes, insulin usage, height, weight and systolic and diastolic blood pressure, which resulted in an average retinal and choroidal thickness increase of 0.13 μm (95%CI: 0.02-0.24 μm, P=0.024) and 0.65 μm (95%CI: 0.13-1.16 μm, P=0.013) for each additional kg of grip strength. Consistent results were obtained in the analyses in ETDRS 9 sectors.

**Conclusion:** Greater hand grip strength was found to be significantly associated with thicker retinal and choroidal layers in diabetic patients. Grip strength may provide a useful indicator of retinal health in diabetic patients. Further studies are needed to determine directionality and causality, and to examine whether improving muscle strength has a positive effect on retinal and choroidal thickness.

**Article summary:** *Strengths and limitations of this study:*

- Grip strength is an indicator of upper limb muscle function and tension, and it is one of the important indicators of aging in the human population. This is one of the few studies to examined the relationship between grip strength and retinal thickness and choroidal thickness in a large sample size of patients with diabetes mellitus.
- Greater hand grip strength was found to be significantly associated with thicker retinal and choroidal thickness in diabetic patients, and grip strength may provide an easily-administered marker of retinal health in diabetic patients.
- Causal inferences could not be inferred due to the cross-sectional design of the study.
- The dynamic changes in grip strength and their impact on retinal and choroidal thickness are warranted to be explored in longitudinal studies.

## Introduction

Diabetes has become a growing health problem, which can cause cardiovascular diseases and death.^1^ Diabetic retinopathy may lead to severe visual disorders. One in every 11 adults in the world has diabetes, and it was estimated that 415 million people between the ages of 20 and 79 had diabetes in 2015, and it is expected to reach 642 million people by 2040.^2^ Among the diabetes patients in this study, 34.6% had diabetic retinopathy (DR), accounting for 2.6% of all blindness.^3 4^ At present, optical coherence tomography (OCT) examinations used in clinics provide an accurate reflection of fundus changes, but widespread performance of this approach is expensive and inconvenient.

Grip strength is an indicator of upper limb muscle function and tension, and it is one of the important indicators of aging in the human population.^5^ Handgrip strength is easy to measure and has great clinical significance. Many studies have reported that low grip strength is associated with type 2 diabetes.^6 7^ Increased grip strength was independently associated with a lower prevalence of prediabetes in Chinese adults.^8^ Fukuda et al.^9^ reported that muscle quality was significantly associated with the presence of DR among patients with type 2 diabetes. Evidence has suggested that the size of a patience’s retinal artery is independently related to grip strength in elderly patients living in Scotland.^10^ The Beaver Dam Eye Study indicated that men with increased handgrip strength had lower odds of early and later age-related macular degeneration by 28% and 55%, respectively.^11^

The retina and choroid are important layers in the eyeball, and retinal and choroidal thickness are both useful indicators of overall eye health.^12^ The choroid provides the retinal pigment epithelium with metabolic support and nourishes the optic nerve, choroid and outer retina. The structure and function of the choroidal vasculature is essential for the retina to function properly.^13^ Abnormalities of retinal and choroidal thickness have been involved in the development and progression of DR. The choroid in patients with diabetes but without DR is thinner than that in healthy people but thicker than that in patients with DR.^14^ Thinning of the choroid is considered to be an early sign of diabetes in patients with no clinical DR.^15^ To the best of our knowledge, the relationship between grip strength and retinal or choroidal thickness has not been explored in previous studies. Therefore, to address this important consideration, the objective of this study was to determine the relationships between grip strength and retinal or choroidal thickness in Chinese patients with type 2 diabetes mellitus using swept-source optical coherence tomography (SS-OCT).

## Methods

### Subjects

This cross-sectional study was performed at the Zhongshan Ophthalmic Centre (ZOC), Sun Yat-sen University, China. The protocol of this study was approved by the Institute Ethics Committee of (ZOC). The study was performed in accordance with the tenets of the Helsinki Declaration. All participants signed a written informed consent form before entering.

Eligible subjects included ocular treatment-naïve patients with type 2 diabetes between the ages of 30 and 80 years old. Participants were excluded if they had any of the following conditions: evidence of diabetic retinopathy based on ETDRS 7 photography, an axial length > 30 mm, a spherical equivalent > −10D, a cylinder degree of 3.0 diopters or more in either eye, anisometropia of 2.0 diopters or more, a C/D ratio ≥ 0.5 or inter-eye asymmetry ≥ 0.2, a history of ocular disease (except for light cataract) or trauma, a history of ocular laser or surgical interventions and/or a history of systemic diseases such as stroke, chronic kidney CKD, cancer or chronic obstructive pulmonary disease (COPD), history of diabetic polyneuropathy.^16^

### Systemic and laboratory tests

Systemic data was collected via standardized questionnaires, including age, sex, length of time patient has had diabetes, medication compliance, presence of other systematic chronic diseases and risk factors. Height, weight, systolic blood pressure (SBP) and diastolic blood pressure (DBP) were also measured. Blood and urine samples were obtained from all patients, and they were analysed for the following parameters: renal profiles that included serum creatinine, microalbuminuria and other parameters, including haemoglobin A1c (HbA1c), total cholesterol (TC), high-density lipoprotein cholesterol (HDL-C), low-density lipoprotein cholesterol (LDL-C) and triglyceride (TG).

### Hand Grip Strength (HGS)

Hand grip strength (HGS) is a proxy of muscle mass and strength. HGS was measured twice for each hand with a dynamometer (Yuejian™ WL-1000, Nantong, China) in kilograms (kg). A trained examiner explained and demonstrated the protocol to each participant. The grip test was performed in the standing position unless the participant was physically limited. The participant’s shoulder was abducted and neutrally rotated, with the elbow at 90° flexion, and the forearm and wrist were in a neutral position. Each participant was randomly directed to begin the test with his or her dominant or nondominant hand and was asked to use one of their hands to squeeze the handle of the dynamometer “as hard as possible for a couple of seconds”. The test was then repeated for the other hand. Each hand was tested twice, alternating hands between tests. The mean score of the four measurements that included both hands was used in the analysis. As prior or present conditions may affect a person’s grip strength, participants that had hand surgery within the past 6 months or severe hand pain or arthritis in the wrists were excluded from the HGS measurement.

### Ocular examination

Comprehensive ocular examinations were conducted for all participants. The anterior and posterior segments were evaluated by slit-lamp biomicroscopy and ophthalmoscopy. The evaluations of visual acuity and intraocular pressure (IOP) were regularly performed. Ocular biometric parameters were obtained using optical low-coherence reflectometry (Lenstar LS900; Haag-Streit AG, Koeniz, Switzerland). If the AL exceeded the measurement range of the Lenstar (32 mm), coherence interferometry (IOL Master; Carl Zeiss Meditec, Oberkochen, Germany) was used instead. Auto refraction was measured by an autorefractor (KR8800; Topcon, Tokyo, Japan) after pupil dilation. The retinal photography was performed in accordance with ETDRS 7 field criteria using a digital fundus camera (Canon CR-2, Tokyo, Japan) after pupil dilation. DR was diagnosed using international clinical diabetic retinopathy and diabetic macular edema disease severity scales.

### SS-OCT Imaging

The SS-OCT (DRI OCT-2 Triton; Topcon, Tokyo, Japan) instrument was used to obtain high-definition retinal and choroidal images. This device has the speed of 100,000 A-scans/s and yields an 8 μm axial resolution in tissue. The method of retinal and choroidal imaging with SS-OCT has been described elsewhere in detail.^17^ Three-dimensional (3-D) imaging scans were obtained using 6×6mm raster scan protocol centred on the macula. The resulting images were analysed with the automated layer segmentation software, which is built into the SS-OCT system. The RT and CT in the nine subfields defined by the ETDRS were automatically calculated and displayed. The ETDRS grid divides the macular into two rings, including the inner and outer rings, which were 1-3 mm and 3-6 mm, respectively. The individual grids are referred to as the centre field, inner superior, inner nasal, inner inferior, inner temporal, outer superior, outer nasal, outer inferior and outer temporal (Figure 1). In addition, the average RT and CT in all nine grids were calculated, and all OCT scans were performed by the same experienced technician who was blind to the study protocol. Before the scan was conducted, it was verified that none of the patients had consumed drinks with caffeine or alcohol or had taken analgesic medications for at least 24 h prior to the procedure. Only subjects with eligible images (i.e., image quality index > 50, without eye movement, without artefacts and without segmentation failure) were included in this study.

**Figure 1.**
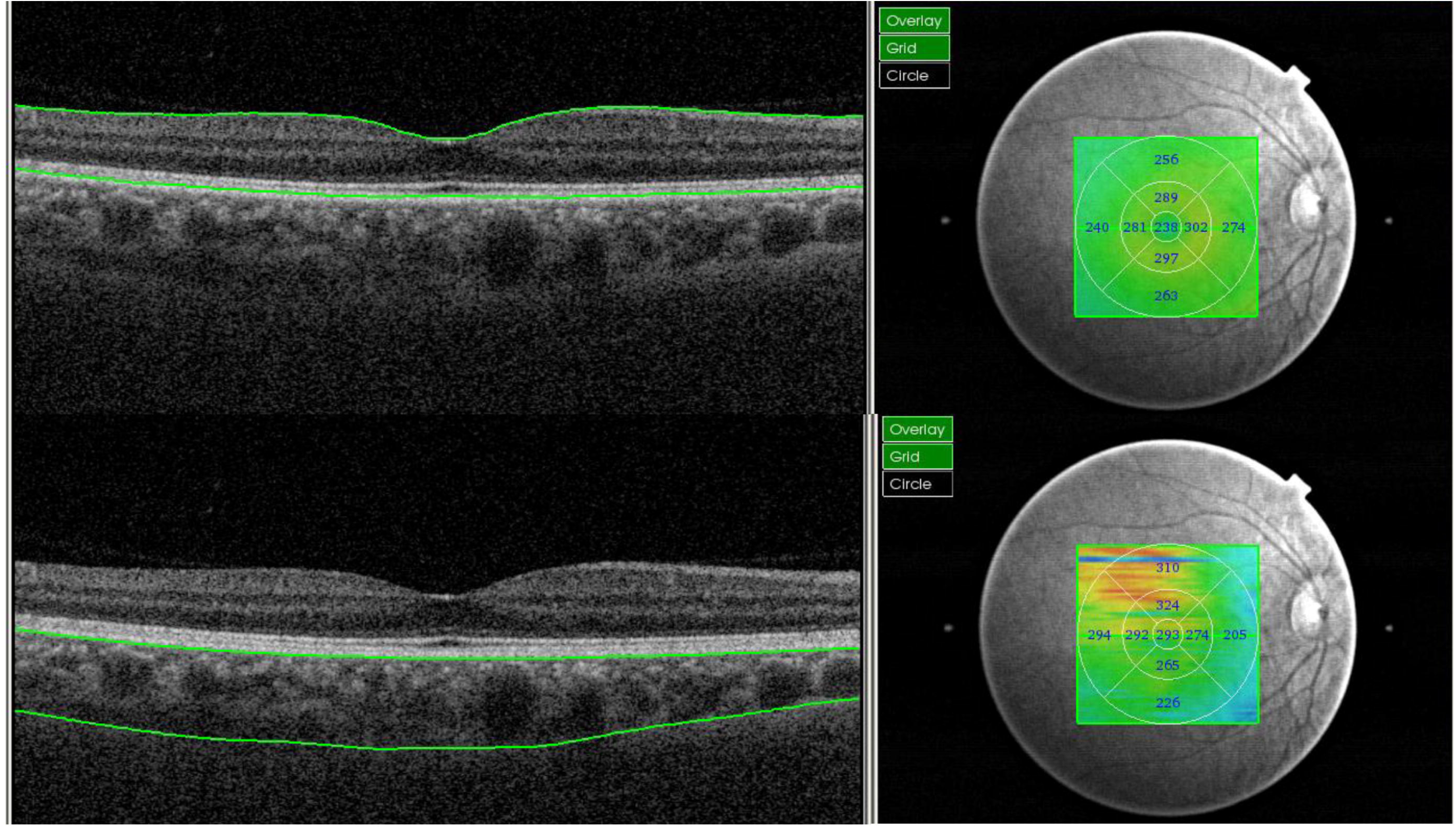
Illustration of measurements of retinal and choroidal thickness in ETDRS 9 sectors using SS-OCT. (A) Retinal thickness; (B) Choroidal thickness.

### Statistical analyses

According to the scores of grip strength from low to high, the participants were divided into four groups: Q1, Q2, Q3 and Q4. The Kolmogorov-Smirnov test conducted to verify the normal distribution. The Fisher’s exact test was used for categorical variables. After normality was confirmed, the ANOVA test was conducted to evaluate the inter-group difference of demographic, systemic and ocular parameters. Bivariate scatter plots were created to display the potential factors affecting CT. Linear regression analyses were performed to assess the association of CT with demographic or ocular parameters, such as age, sex, AL, HbA1c and other parameters previously mentioned.^17^ Univariate analysis showed that the predictive variables were significant, which were then entered into the final multivariate equation. A *P* value <0.05 was considered statistically significant. All analyses were performed using Stata, version 14.0 (Stata Corporation, College Station, TX, USA).

## Results

A total of 1,029 patients without any evidence of DR were included in the present study, which included 616 (59.86%) female with a mean age of 64.5±7.7 years. Table 1 presents the characteristics of the participants. Differences in gender, age, height, weight, total cholesterol, serum creatinine, HDL-C, CCT and axial length between each group were found to be statistically significant (*P* < 0.05).

**Table 1.** Demographic and clinical characteristics of diabetic patients without retinopathy.

| Characteristics | All | Quarter of grip strength |  |  |  | P-value |
| --- | --- | --- | --- | --- | --- | --- |
|  |  | Q1 (Lowest) | Q2 | Q3 | Q4 (Highest) |  |
| No. of subjects | 1029 | 258 | 260 | 255 | 256 | - |
| Female, % | 616(59.86%) | 224(86.82%) | 215(82.69%) | 129(50.59%) | 48(18.75%) | **<0.001** |
| Use of insulin, % | 163(16.03%) | 40(15.69%) | 39(15.18%) | 39(15.42%) | 45(17.86%) | 0.836 |
| Age, year | 64.5±7.7 | 66.2±7.2 | 64.7±7.4 | 63.6±7.8 | 63.6±8.1 | **<0.001** |
| Duration of diabetes, year | 8.1±6.6 | 8.7±6.7 | 8.3±6.7 | 7.5±6.3 | 8.1±6.6 | 0.214 |
| HbA1c, % | 6.8±1.3 | 6.8±1.2 | 6.7±1.3 | 6.8±1.3 | 6.9±1.4 | 0.507 |
| Height, cm | 160.1±8.1 | 155.4±6.9 | 157.8±6.7 | 161.7±6.9 | 165.5±8.0 | **<0.001** |
| Weight, kg | 63.7±11.5 | 59.5±9.9 | 60.7±9.9 | 65.0±10.0 | 69.7±13.0 | **<0.001** |
| Systolic blood pressure, mm Hg | 133.7±18.5 | 132.6±19.2 | 134.3±18.4 | 134.1±18.2 | 133.9±18.1 | 0.714 |
| Diastolic blood pressure, mm Hg | 70.7±10.4 | 69.6±10.8 | 70.3±9.9 | 71.4±10.0 | 71.4±10.9 | 0.112 |
| Total cholesterol, mmol/L | 4.8±1.0 | 4.9±1.0 | 5.0±1.1 | 4.8±1.0 | 4.6±1.0 | **0.001** |
| Serum creatinine, µmol/L | 70.3±19.3 | 68.0±20.2 | 65.2±19.8 | 71.3±18.3 | 76.6±17.0 | **<0.001** |
| HDL-c, mmol/L | 1.3±0.4 | 1.4±0.4 | 1.4±0.4 | 1.3±0.4 | 1.2±0.4 | **<0.001** |
| LDL-c, mmol/L | 3.1±0.9 | 3.1±0.9 | 3.2±1.0 | 3.1±0.9 | 3.0±0.9 | 0.122 |
| Triglycerides, mmol/L | 2.3±1.6 | 2.4±1.6 | 2.3±1.5 | 2.3±1.7 | 2.3±1.7 | 0.976 |
| C-reactive protein, mg/L | 2.7±7.4 | 2.8±3.5 | 3.1±12.3 | 2.4±3.8 | 2.4±6.5 | 0.658 |
| Microalbuminuria, mg/mL | 4.1±14.9 | 4.7±14.7 | 4.9±23.0 | 3.3±8.0 | 3.6±9.0 | 0.530 |
| BCVA, logMAR | 0.2±0.1 | 0.2±0.1 | 0.2±0.1 | 0.2±0.2 | 0.2±0.1 | 0.133 |
| Intraocular pressure, mmHg | 16.2±2.7 | 16.4±2.8 | 16.2±2.6 | 15.9±2.7 | 16.1±2.8 | 0.278 |
| Central corneal thickness, µm | 546.1±31.5 | 545.8±29.6 | 544.8±30.8 | 542.8±33.1 | 550.9±31.9 | **0.026** |
| Axial length, mm | 23.6±1.1 | 23.4±1.0 | 23.5±1.2 | 23.6±1.1 | 23.8±1.0 | **<0.001** |

BCVA=best corrected visual acuity. Bold indicates statistically significant.

The average retinal and choroidal thicknesses for all participants were 274.4±15.3 μm and 184.5±73.6 μm, respectively. Table 2 presents the distribution of retinal thickness and choroidal thickness in each sector across groups. There were statistically significant differences in retinal and choroidal thicknesses among the four groups (P < 0.001, retinal thickness in the outer inferior P=0.0483). Both retinal and choroidal thicknesses increased in the higher quartile of grip strength (Figure 2).

**Table 2.** Differences of retinal and choroidal thickness among groups by quartiles of grips strength.

| Parameters | All | Quartile 1 | Quartile 2 | Quartile 3 | Quartile 4 | P-value |
| --- | --- | --- | --- | --- | --- | --- |
| **Retinal thickness** |  |  |  |  |  |  |
| Outer Superior, $\mu\text{m}$ | 265.8±18.9 | 261.8±15.6 | 265.9±20.0 | 266.6±22.7 | 268.8±15.9 | **<0.001** |
| Inner Superior, $\mu\text{m}$ | 301.5±18.3 | 296.6±15.1 | 301.3±19.1 | 302.3±19.7 | 305.9±18.1 | **<0.001** |
| Outer Temporal, $\mu\text{m}$ | 251.5±16.9 | 247.4±15.1 | 250.5±18.2 | 252.6±17.1 | 255.5±15.8 | **<0.001** |
| Inner Temporal, $\mu\text{m}$ | 288.8±17.7 | 284.4±15.8 | 286.8±17.2 | 290.2±18.8 | 294.0±17.6 | **<0.001** |
| Central field, $\mu\text{m}$ | 228.4±23.5 | 222.5±19.1 | 225.6±22.0 | 231.6±25.7 | 234.2±25.0 | **<0.001** |
| Inner Nasal, $\mu\text{m}$ | 300.9±18.4 | 296.4±16.1 | 299.6±18.9 | 301.9±18.5 | 305.6±18.8 | **<0.001** |
| Outer Nasal, $\mu\text{m}$ | 280.0±17.9 | 276.3±15.8 | 279.2±18.7 | 280.6±18.6 | 284.0±17.6 | **<0.001** |
| Inner Inferior, $\mu\text{m}$ | 297.2±19.6 | 292.8±17.2 | 295.8±20.5 | 297.9±19.7 | 302.5±19.8 | **<0.001** |
| Outer Inferior, $\mu\text{m}$ | 255.8±19.0 | 253.1±15.1 | 255.7±24.3 | 256.6±18.1 | 257.6±17.0 | **0.0483** |
| Average, $\mu\text{m}$ | 274.4±15.3 | 270.2±12.9 | 273.4±15.6 | 275.6±16.1 | 278.7±15.1 | **<0.001** |
| **Choroidal thickness** |  |  |  |  |  |  |
| Outer Superior, $\mu\text{m}$ | 192.5±75.1 | 178.2±74.2 | 188.9±74.1 | 193.7±74.0 | 209.4±75.3 | **<0.001** |
| Inner Superior, $\mu\text{m}$ | 202.0±81.5 | 187.2±81.1 | 200.2±79.5 | 201.1±82.1 | 219.7±80.2 | **<0.001** |
| Outer Temporal, $\mu\text{m}$ | 173.9±65.0 | 159.1±59.6 | 177.0±62.8 | 170.6±67.9 | 189.0±66.2 | **<0.001** |
| Inner Temporal, $\mu\text{m}$ | 195.4±74.4 | 179.3±69.1 | 199.1±72.5 | 191.1±77.3 | 212.3±74.9 | **<0.001** |
| Central field, $\mu\text{m}$ | 201.9±83.8 | 184.9±80.4 | 202.6±83.2 | 199.1±85.8 | 221.0±82.4 | **<0.001** |
| Inner Nasal, $\mu\text{m}$ | 189.6±85.5 | 171.4±81.0 | 190.6±85.0 | 189.5±86.5 | 207.2±86.1 | **<0.001** |
| Outer Nasal, $\mu\text{m}$ | 149.9±79.5 | 132.9±72.0 | 151.5±80.4 | 153.3±81.5 | 162.0±81.4 | **<0.001** |
| Inner Inferior, $\mu\text{m}$ | 188.2±83.5 | 166.4±77.2 | 190.5±83.6 | 187.6±83.4 | 208.6±84.7 | **<0.001** |
| Outer Inferior, $\mu\text{m}$ | 167.0±78.1 | 147.0±70.2 | 169.1±78.5 | 166.8±79.3 | 185.3±79.7 | **<0.001** |
| Average, $\mu\text{m}$ | 184.5 ±73.6 | 167.4 ±69.1 | 185.5 ±72.9 | 183.6 ±74.8 | 201.6 ±73.8 | **<0.001** |

Bold indicates statistically significant.

**Figure 2.**
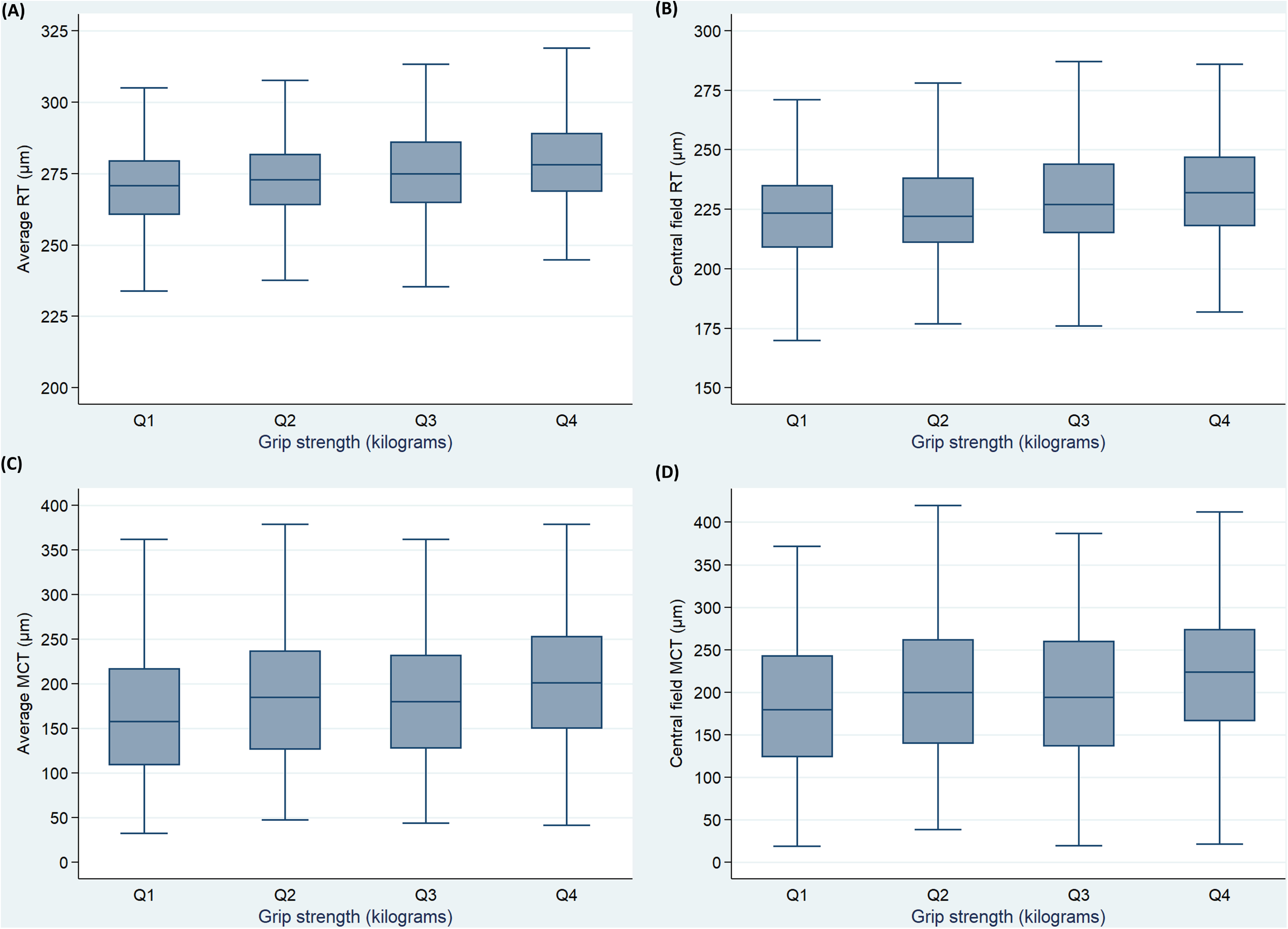
Boxplots showing the distribution of retinal and choroidal thickness stratified by quartiles of the handgrip strength. (A) (A)Average retinal thickness versus grip strength; (B) Retinal thickness in central field versus grip strength; (C) Average choroidal thickness versus grip strength; (D) Choroidal thickness in central field versus grip strength.

Table 3 presents the association between grips strength and retinal thickness based on multivariable regression analyses. After adjusting for age and gender, the average retinal and choroidal thickness increased by 0.14 μm (P=0.011) and 0.57 μm (P=0.037) for each kg of increased grip strength. There was a statistically significant association between grip strength scores and retinal thickness in at least one group in each region except the outer inferior. After making adjustments for age, sex, axial length, HbA1c, duration of diabetes, insulin usage, height, weight and systolic and diastolic blood pressure, the average retinal and choroidal thickness increased by 0.13 μm (*P*=0.024) and 0.65 μm (*P*=0.013) for each kg of increased grip strength. There was a statistically significant association between grip strength scores and retinal thickness in at least one group in each region except the outer inferior, the inner inferior, nasal and temporal.

**Table 3.** Association of grip strength and retinal thickness by multivariate linear regression analyses.

| Retinal thickness | Model 1* |  |  |  | Model 2† |  |  |
| --- | --- | --- | --- | --- | --- | --- | --- |
|  | Coefficient | 95%CI | P-value |  | Coefficient | 95%CI | P-value |
| **Average, $\mu\text{m}$** |  |  |  |  |  |  |  |
| Quantile 1 | 1.00 (Ref) |  |  |  | 1.00 (Ref) |  |  |
| Quantile 2 | 2.16 | (-0.28 to 4.61) | 0.083 |  | 1.71 | (-0.75 to 4.168) | 0.174 |
| Quantile 3 | 2.10 | (-0.48 to 4.69) | 0.111 |  | 1.33 | (-1.28 to 3.935) | 0.319 |
| Quantile 4 | 3.68 | (0.81 to 6.55) | **0.012** |  | 3.35 | (0.44 to 6.256) | **0.024** |
| Per 1-kg increase | 0.14 | (0.03 to 0.25) | **0.011** |  | 0.13 | (0.02 to 0.24) | **0.024** |
| **Central field, $\mu\text{m}$** |  |  |  |  |  |  |  |
| Quantile 1 | 1.00 (Ref) |  |  |  | 1.00 (Ref) |  |  |
| Quantile 2 | 2.50 | (-1.42 to 6.43) | 0.211 |  | 1.90 | (-2.04 to 5.831) | 0.345 |
| Quantile 3 | 5.08 | (0.92 to 9.24) | **0.017** |  | 4.62 | (0.45 to 8.790) | **0.030** |
| Quantile 4 | 4.53 | (-0.08 to 9.13) | 0.054 |  | 4.15 | (-0.49 to 8.801) | 0.080 |
| Per 1-kg increase | 0.16 | (-0.02 to 0.33) | 0.082 |  | 0.15 | (-0.03 to 0.32) | 0.109 |
| **Outer Superior, $\mu\text{m}$** |  |  |  |  |  |  |  |
| Quantile 1 | 1.00 (Ref) |  |  |  | 1.00 (Ref) |  |  |
| Quantile 2 | 3.03 | (-0.08 to 6.14) | 0.056 |  | 2.65 | (-0.43 to 5.725) | 0.091 |
| Quantile 3 | 2.49 | (-0.80 to 5.78) | 0.138 |  | 1.47 | (-1.79 to 4.734) | 0.375 |
| Quantile 4 | 4.44 | (0.80 to 8.08) | **0.017** |  | 3.74 | (0.11 to 7.373) | **0.043** |
| Per 1-kg increase | 0.19 | (0.05 to 0.33) | **0.008** |  | 0.16 | (0.02 to 0.30) | **0.022** |
| **Outer Inferior, $\mu\text{m}$** |  |  |  |  |  |  |  |
| Quantile 1 | 1.00 (Ref) |  |  |  | 1.00 (Ref) |  |  |
| Quantile 2 | 1.49 | (-1.65 to 4.62) | 0.352 |  | 1.27 | (-1.83 to 4.373) | 0.420 |
| Quantile 3 | 1.17 | (-2.15 to 4.49) | 0.489 |  | 0.49 | (-2.79 to 3.778) | 0.768 |
| Quantile 4 | 1.86 | (-1.82 to 5.53) | 0.321 |  | 1.90 | (-1.76 to 5.561) | 0.308 |
| Per 1-kg increase | 0.06 | (-0.08 to 0.21) | 0.368 |  | 0.06 | (-0.08 to 0.20) | 0.377 |
| **Outer Nasal, $\mu\text{m}$** |  |  |  |  |  |  |  |
| Quantile 1 | 1.00 (Ref) |  |  |  | 1.00 (Ref) |  |  |
| Quantile 2 | 1.72 | (-1.16 to 4.60) | 0.243 |  | 1.63 | (-1.20 to 4.475) | 0.259 |
| Quantile 3 | 1.16 | (-1.89 to 4.21) | 0.457 |  | 0.55 | (-2.46 to 3.557) | 0.721 |
| Quantile 4 | 3.64 | (0.26 to 7.02) | **0.035** |  | 3.46 | (0.11 to 6.810) | **0.043** |
| Per 1-kg increase | 0.14 | (0.01 to 0.27) | **0.031** |  | 0.14 | (0.01 to 0.26) | **0.040** |
| **Outer Temporal, $\mu\text{m}$** |  |  |  |  |  |  |  |
| Quantile 1 | 1.00 (Ref) |  |  |  | 1.00 (Ref) |  |  |
| Quantile 2 | 1.94 | (-0.78 to 4.66) | 0.162 |  | 1.83 | (-0.80 to 4.449) | 0.172 |
| Quantile 3 | 1.88 | (-1.01 to 4.76) | 0.202 |  | 1.02 | (-1.76 to 3.799) | 0.472 |
| Quantile 4 | 3.36 | (0.17 to 6.56) | **0.039** |  | 2.72 | (-0.37 to 5.818) | 0.085 |
| Per 1-kg increase | 0.14 | (0.02 to 0.27) | **0.022** |  | 0.12 | (0.00 to 0.24) | 0.053 |
| **Inner Superior, $\mu\text{m}$** |  |  |  |  |  |  |  |
| Quantile 1 | 1.00 (Ref) |  |  |  | 1.00 (Ref) |  |  |
| Quantile 2 | 3.62 | (0.61 to 6.63) | **0.018** |  | 3.18 | (0.13 to 6.226) | **0.041** |
| Quantile 3 | 2.39 | (-0.80 to 5.58) | 0.142 |  | 1.68 | (-1.54 to 4.912) | 0.306 |
| Quantile 4 | 4.52 | (0.99 to 8.05) | **0.012** |  | 4.46 | (0.86 to 8.053) | **0.015** |
| Per 1-kg increase | 0.15 | (0.01 to 0.29) | **0.032** |  | 0.14 | (0.00 to 0.28) | **0.046** |
| **Inner Inferior, $\mu\text{m}$** |  |  |  |  |  |  |  |
| Quantile 1 | 1.00 (Ref) |  |  |  | 1.00 (Ref) |  |  |
| Quantile 2 | 1.73 | (-1.47 to 4.94) | 0.289 |  | 0.88 | (-2.29 to 4.059) | 0.586 |
| Quantile 3 | 1.01 | (-2.39 to 4.40) | 0.561 |  | 0.01 | (-3.36 to 3.377) | 0.995 |
| Quantile 4 | 3.51 | (-0.25 to 7.27) | 0.067 |  | 3.38 | (-0.37 to 7.133) | 0.077 |
| Per 1-kg increase | 0.15 | (0.00 to 0.29) | **0.048** |  | 0.13 | (-0.01 to 0.28) | 0.069 |
| **Inner Nasal, $\mu\text{m}$** |  |  |  |  |  |  |  |
| Quantile 1 | 1.00 (Ref) |  |  |  | 1.00 (Ref) |  |  |
| Quantile 2 | 2.15 | (-0.89 to 5.18) | 0.165 |  | 1.40 | (-1.67 to 4.473) | 0.372 |
| Quantile 3 | 2.02 | (-1.19 to 5.24) | 0.217 |  | 1.25 | (-2.00 to 4.509) | 0.451 |
| Quantile 4 | 3.86 | (0.30 to 7.42) | **0.034** |  | 3.50 | (-0.13 to 7.124) | 0.059 |
| Per 1-kg increase | 0.15 | (0.01 to 0.28) | **0.038** |  | 0.13 | (-0.01 to 0.27) | 0.067 |
| **Inner Temporal, $\mu\text{m}$** |  |  |  |  |  |  |  |
| Quantile 1 | 1.00 (Ref) |  |  |  | 1.00 (Ref) |  |  |
| Quantile 2 | 1.34 | (-1.55 to 4.23) | 0.363 |  | 0.66 | (-2.27 to 3.577) | 0.660 |
| Quantile 3 | 1.82 | (-1.24 to 4.88) | 0.243 |  | 0.91 | (-2.18 to 4.008) | 0.563 |
| Quantile 4 | 3.40 | (0.02 to 6.79) | **0.049** |  | 2.84 | (-0.61 to 6.287) | 0.106 |
| Per 1-kg increase | 0.15 | (0.02 to 0.28) | **0.021** |  | 0.13 | (0.00 to 0.26) | 0.055 |

Model 1: adjusted for age and sex.
Model 2: adjusted for age, gender, axial length, HbA1c, diabetes duration, use of insulin, height, weight, systolic blood pressure, and diastolic blood pressure.
Bold indicates statistically significant.

There was a statistically significant correlation between grip strength and choroidal thickness in at least one group in each region, except in the upper and inner superior regions. There was a statistically significant correlation between grip strength and choroidal thickness in at least one group in each region, except in the outer superior region (Table 4).

**Table 4.** Association of grip strength and choroidal thickness by multivariate linear regression analyses.

| Choroidal thickness | Model 1* |  |  |  | Model 2† |  |  |
| --- | --- | --- | --- | --- | --- | --- | --- |
|  | Coefficient | 95%CI | P-value |  | Coefficient | 95%CI | P-value |
| **Average, $\mu\text{m}$** |  |  |  |  |  |  |  |
| Quantile 1 |  | 1.00 (Ref) |  |  |  | 1.00 (Ref) |  |
| Quantile 2 | 13.37 | (1.43 to 25.31) | **0.028** |  | 13.51 | (2.26 to 24.75) | **0.019** |
| Quantile 3 | 3.66 | (-8.98 to 16.31) | 0.570 |  | 1.66 | (-10.26 to 13.58) | 0.784 |
| Quantile 4 | 17.52 | (3.52 to 31.51) | **0.014** |  | 20.42 | (7.14 to 33.69) | **0.003** |
| Per 1-kg increase | 0.57 | (0.03 to 1.11) | **0.037** |  | 0.65 | (0.13 to 1.16) | **0.013** |
| **Central field, $\mu\text{m}$** |  |  |  |  |  |  |  |
| Quantile 1 |  | 1.00 (Ref) |  |  |  | 1.00 (Ref) |  |
| Quantile 2 | 13.00 | (-0.82 to 26.83) | 0.065 |  | 13.47 | (0.50 to 26.45) | **0.042** |
| Quantile 3 | 1.02 | (-13.63 to 15.66) | 0.892 |  | -1.39 | (-15.14 to 12.35) | 0.842 |
| Quantile 4 | 18.06 | (1.85 to 34.27) | **0.029** |  | 21.39 | (6.08 to 36.70) | **0.006** |
| Per 1-kg increase | 0.57 | (-0.05 to 1.20) | 0.072 |  | 0.65 | (0.06 to 1.24) | **0.031** |
| **Outer Superior, $\mu\text{m}$** |  |  |  |  |  |  |  |
| Quantile 1 |  | 1.00 (Ref) |  |  |  | 1.00 (Ref) |  |
| Quantile 2 | 5.65 | (-6.56 to 17.87) | 0.364 |  | 5.01 | (-6.78 to 16.80) | 0.405 |
| Quantile 3 | 0.89 | (-12.05 to 13.83) | 0.893 |  | -1.34 | (-13.84 to 11.15) | 0.833 |
| Quantile 4 | 10.78 | (-3.54 to 25.11) | 0.140 |  | 13.60 | (-0.32 to 27.51) | 0.055 |
| Per 1-kg increase | 0.39 | (-0.16 to 0.94) | 0.167 |  | 0.47 | (-0.07 to 1.00) | 0.088 |
| **Outer Inferior, $\mu\text{m}$** |  |  |  |  |  |  |  |
| Quantile 1 |  | 1.00 (Ref) |  |  |  | 1.00 (Ref) |  |
| Quantile 2 | 16.85 | (4.29 to 29.40) | **0.009** |  | 16.59 | (4.53 to 28.65) | **0.007** |
| Quantile 3 | 7.11 | (-6.19 to 20.40) | 0.295 |  | 4.83 | (-7.95 to 17.61) | 0.459 |
| Quantile 4 | 22.49 | (7.77 to 37.21) | **0.003** |  | 24.36 | (10.12 to 38.59) | **0.001** |
| Per 1-kg increase | 0.73 | (0.16 to 1.30) | **0.012** |  | 0.77 | (0.22 to 1.32) | **0.006** |
| **Outer Nasal, $\mu\text{m}$** |  |  |  |  |  |  |  |
| Quantile 1 |  | 1.00 (Ref) |  |  |  | 1.00 (Ref) |  |
| Quantile 2 | 13.79 | (0.77 to 26.81) | **0.038** |  | 14.83 | (2.52 to 27.14) | **0.018** |
| Quantile 3 | 8.24 | (-5.55 to 22.03) | 0.241 |  | 7.26 | (-5.79 to 20.30) | 0.275 |
| Quantile 4 | 13.45 | (-1.82 to 28.71) | 0.084 |  | 16.76 | (2.23 to 31.28) | **0.024** |
| Per 1-kg increase | 0.38 | (-0.21 to 0.97) | 0.202 |  | 0.47 | (-0.09 to 1.03) | 0.102 |
| **Outer Temporal, $\mu\text{m}$** |  |  |  |  |  |  |  |
| Quantile 1 |  | 1.00 (Ref) |  |  |  | 1.00 (Ref) |  |
| Quantile 2 | 14.00 | (3.40 to 24.60) | **0.010** |  | 13.19 | (2.87 to 23.50) | **0.012** |
| Quantile 3 | 2.00 | (-9.23 to 13.22) | 0.727 |  | -0.18 | (-11.12 to 10.75) | 0.974 |
| Quantile 4 | 17.94 | (5.51 to 30.36) | **0.005** |  | 20.07 | (7.90 to 32.24) | **0.001** |
| Per 1-kg increase | 0.64 | (0.16 to 1.12) | **0.009** |  | 0.71 | (0.24 to 1.18) | **0.003** |
| **Inner Superior, $\mu\text{m}$** |  |  |  |  |  |  |  |
| Quantile 1 |  | 1.00 (Ref) |  |  |  | 1.00 (Ref) |  |
| Quantile 2 | 8.10 | (-5.28 to 21.48) | 0.235 |  | 8.11 | (-4.63 to 20.85) | 0.212 |
| Quantile 3 | 0.04 | (-14.13 to 14.21) | 0.996 |  | -2.23 | (-15.73 to 11.28) | 0.746 |
| Quantile 4 | 13.39 | (-2.29 to 29.08) | 0.094 |  | 16.60 | (1.56 to 31.64) | **0.031** |
| Per 1-kg increase | 0.48 | (-0.13 to 1.08) | 0.122 |  | 0.55 | (-0.03 to 1.13) | 0.062 |
| **Inner Inferior, $\mu\text{m}$** |  |  |  |  |  |  |  |
| Quantile 1 |  | 1.00 (Ref) |  |  |  | 1.00 (Ref) |  |
| Quantile 2 | 19.10 | (5.49 to 32.72) | **0.006** |  | 19.74 | (6.77 to 32.71) | **0.003** |
| Quantile 3 | 8.81 | (-5.61 to 23.22) | 0.231 |  | 7.25 | (-6.49 to 20.99) | 0.301 |
| Quantile 4 | 26.26 | (10.30 to 42.22) | **0.001** |  | 29.10 | (13.80 to 44.41) | **0.000** |
| Per 1-kg increase | 0.85 | (0.24 to 1.47) | **0.007** |  | 0.92 | (0.33 to 1.52) | **0.002** |
| **Inner Nasal, $\mu\text{m}$** |  |  |  |  |  |  |  |
| Quantile 1 |  | 1.00 (Ref) |  |  |  | 1.00 (Ref) |  |
| Quantile 2 | 13.98 | (-0.03 to 27.99) | 0.051 |  | 14.48 | (1.29 to 27.67) | **0.031** |
| Quantile 3 | 4.34 | (-10.50 to 19.18) | 0.566 |  | 2.01 | (-11.97 to 15.98) | 0.778 |
| Quantile 4 | 17.43 | (1.00 to 33.86) | **0.038** |  | 20.37 | (4.80 to 35.94) | **0.010** |
| Per 1-kg increase | 0.52 | (-0.11 to 1.15) | 0.107 |  | 0.58 | (-0.02 to 1.18) | 0.058 |
| **Inner Temporal, $\mu\text{m}$** |  |  |  |  |  |  |  |
| Quantile 1 |  | 1.00 (Ref) |  |  |  | 1.00 (Ref) |  |
| Quantile 2 | 15.67 | (3.43 to 27.91) | **0.012** |  | 15.97 | (4.32 to 27.62) | **0.007** |
| Quantile 3 | 0.55 | (-12.41 to 13.51) | 0.934 |  | -1.20 | (-13.55 to 11.14) | 0.848 |
| Quantile 4 | 17.77 | (3.42 to 32.12) | **0.015** |  | 21.40 | (7.65 to 35.15) | **0.002** |
| Per 1-kg increase | 0.60 | (0.04 to 1.15) | **0.034** |  | 0.71 | (0.17 to 1.24) | **0.009** |

Model 1: adjusted for age and sex.
Bold indicates statistically significant.

## Discussion

This cross-sectional study reported the relationship existing between handgrip strength and diabetes, analyzing retinal and choroidal thickness. Authors stratified the grip strength results in 4 groups and all the analysis were conducted explaining the differences between these groups that included a large population with aged between 30 and 80. We found significant differences in retinal thickness and choroidal thickness among people with different grip strengths. Multivariate analysis suggested that grip strength in diabetic patients without DR was strongly significant associated with retinal thickness and choroidal thickness, independent of age, gender, axial length, HbA1c, diabetes duration, use of insulin, height, weight, systolic blood pressure and diastolic blood pressure.

The results revealed there was an association between grip strength and retinal or choroidal thickness. Handgrip strength has been shown to be an indicator of aging.^18^ A decrease in grip strength associated with aging correlates with the development of age-related disorders.^19^ Multiple studies have explored the association between handgrip strength and cardiovascular diseases, cognition and all-cause mortality.^5 6 20-23^ In addition to age, the results showed that grip strength is also associated with gender, height and weight.^24^ Therefore, these factors were adjusted in the analysis. The results showed that grip strength was correlated with retinal or choroid thickness in 9 ETDRS domains. The lower the grip strength score was, the thinner the retina and choroid tended to be. The thinning of the retina and choroid can seriously affect the visual function in patients, and it can even lead to blindness.^25^

Lower grip strength scores were associated with thinner retinal or choroidal layers. A possible explanation for this may be related to the blood vessels linking them. The retina and choroid contain a significant number of blood vessels, especially the choroid.^26^ Their thickness often varies greatly depending on the filling state of the blood vessels. Blood vessels are centres for transporting oxygen and nutrients. Thinning of the retina and choroid indicates a decrease in the density and velocity of blood, thus affecting the delivery of oxygen and nutrients to muscles.^27^ It leads to a weakening of muscle strength, which is reflected in the decline in a patient’s grip strength score. In addition, diabetic patients had accelerated aging process, which leads to loss of muscle strength. The presence and severity of diabetic neuropathy was related to gradual loss of muscle strength, and also related to retinal and choroidal thickness. Diabetic nephropathy contributed to loss of muscle strength, and altered retinal and choroidal thickness is a hallmark of renal function in diabetic patients.

This finding has important implications for the development of earlier diagnosis methods for diabetic retinal and choroidal changes. Compared with the OCT examination, handgrip strength measurement is simple and inexpensive, which makes it an economically viable method used in the early evaluation of the progress of fundus lesions in diabetic patients and to monitor their health status.^23 28^ It can be used to implement effective intervention earlier and to improve treatments designed to better control diseases in diabetic patients. Follow-up studies may explore whether grip strength training can slow down the progression of retinal and choroidal thinning. Moreover, while grip strength may provide a useful indicator of retinal health in diabetic patients, further studies are needed to determine directionality and causality, and examining whether improving muscle strength has an influence on retinal and choroidal thickness is also important.

As stated previously, to the best of our knowledge, the present study is the first to identify an association between grip strength and retinal or choroidal thickness. It is a new direction for future research on grip strength. In addition, the subjects covered a wide range of ages, which differs from most previous studies on grip strength that focused on the elderly. The subjects in this study ranged from 30-80 years old, with an average age of 64.5±7.7 years. Moreover, the handgrip strength score was measured by the Jamar J00105 hydraulic manual dynamometer, which has been recommended as a standard tool for handgrip strength testing by the American Society of Hand Therapy (ASHT) and is considered to be the gold standard for handgrip strength testing. The thickness of the retina and choroid was obtained by the same experienced technician using the latest SS-OCT to ensure the results are accurate and reliable. However, further study with larger sample sizes are needed to confirm our findings.

There are several limitations associated with this result. First, this study is a cross-sectional study, which prevents cause-effect inference. In addition, the dynamic changes in grip strength and their impact on retinal and choroidal thickness were not evaluated. Secondly, investigators have just begun to explore the association of grip strength and retinal or choroidal thickness, and more information is needed to gain a better understanding of the connection between them. This study did not explore the long-term impact of grip strength changes on retinal and choroidal thickness or track the progress of diabetes and ocular diseases in each patient. Finally, only patients without DR were included. The generalizability of the findings was therefore reduced.

## Conclusions

In summary, there was significant association between grip strength and retinal and choroidal thickness. Greater hand grip strength was found to be significantly associated with thicker retinal and choroidal thickness in diabetic patients, and grip strength may provide an easily-administered marker of retinal health in diabetic patients. Further studies are needed to determine the directionality and causality, and such studies must examine whether improving muscle strength has an effect on retinal and choroidal thickness.

## Data Availability

Data are available upon reasonable request.

## Acknowledgements

None

## Contributors

Design and conduct of the study (WW, YL, WH); Collection, management, analysis and interpretation of the data (ZQ, WW, YT, MH, LW, YL, WH); Preparation of the manuscript (ZQ, WW, WH); Review and final approval of the manuscript (all authors).

## Patient consent for publication

Not required.

## Ethics approval

The protocol was approved by the Committee of Zhongshan Ophthalmic Center (protocol number IRB#2017-05-0621).

## Provenance and peer review

Not commissioned; externally peer reviewed.

## Data availability statement

Data are available upon reasonable request.

